# Development of Alzheimer’s Disease Risk Score for Future Primary Care: A White-Box Approach

**DOI:** 10.1101/2024.08.02.24311399

**Authors:** Yumiko Wiranto, Devin R Setiawan, Amber Watts, Arian Ashourvan, the Alzheimer’s Disease Neuroimaging Initiative

## Abstract

**Importance:** Interpretable scoring system can contribute to bridge the gap between the timeliness and complexity of diagnosing Alzheimer’s disease (AD) and promote early intervention at non-specialist settings.

**Objective:** To develop a risk score to predict the likelihood of AD with interpretable machine learning using variables that are obtainable at integrated primary care settings.

**Design:** A secondary data analysis including cohort studies from the Alzheimer’s Disease Neuroimaging Initiative (ADNI) and the National Alzheimer’s Coordinating Center (NACC) extracted in August 2023 and March 2024.

**Setting:** The ADNI and NACC are multi-site cohort studies in North America.

**Participants:** Participants with normal cognition or mild cognitive impairment at baseline visit were identified. Participants with the same diagnosis overtime were assigned to the stable group, and those converted to AD were placed in the progressive group.

**Main Outcome(s) and Measure(s):** Cognitive tests and daily functioning measured with Functional Assessment Questionnaire (FAQ) at baseline visit.

**Results:** A total of 676 participants from ADNI and 4592 participants from NACC were identified. After removing incomplete data, 665 ADNI (mean age [SD]: 73.44 [6.90]; 293 [44.1%] female; 374 stable and 291 progressive) and 3657 NACC participants (mean age [SD]: 70.96 [10.03]; 2405 [65.8%] female; 2445 stable and 1212 progressive) remained. Combinations of 4 measures were selected to generate 10 scorecards using FasterRisk algorithm, showing strong performance (area under the curve [AUC] = 0.868-0.892) in ADNI and remaining robust when validated in NACC (AUC = 0.795). The features were Category Animal ≤ 20 (2 points), Trail Making Test B ≤ 143 (−3 points), Logical Memory Delayed ≤ 3 (4 points), Logical Memory Delayed ≤ 8 (3 points), and FAQ ≤ 2 (−5 points). The probable AD risk corresponded to total points: 7.4% (–8), 25.3% (–4), 50% (–1), 74.7% (2), and > 90% (≥ 6). We refer to this model as the (F)unctioning, (LA)nguage, (M)emory, and (E)xecutive functioning or FLAME scorecard.

**Conclusions and Relevance:** Our findings highlight the potential to predict AD development using obtainable information, allowing for applicability at integrated primary care. While our scope centers on AD, this foundation paves the way for other dementia types

**Key Points:** *Question:* Can accessible information, such as demographics, cognitive tests, and functioning questionnaire, yield in reliable results for predicting Alzheimer’s disease development using interpretable machine learning?

*Findings:* The results of 665 participants from the Alzheimer’s Disease Neuroimaging Initiative demonstrated robust performance of determining Alzheimer’s disease development using four separate measures of (F)unctioning, (LA)nguage, (M)emory, and (E)xecutive functioning or the FLAME scorecard. It remains reliable when externally validated with a separate dataset of 3657 participants from the National Alzheimer’s Coordinating Center.

*Meaning:* The FLAME scorecard shows potential to be implemented in integrated primary care settings to promote early detection and intervention of cognitive decline due to Alzheimer’s disease.

## Introduction

As the prevalence of Alzheimer’s disease (AD) continues to rise, timely early detection becomes increasingly urgent. One potential barrier to timely diagnosis is the initial point of contact for many patients: their primary care physicians (PCPs). When individuals first notice memory-related issues, the first healthcare professional they typically go to is their PCPs.

However, many PCPs may not feel confident in delivering a conclusive diagnosis, resulting in delays in obtaining appropriate care.^1,2^ Additionally, even if PCPs refer their patients for further evaluations, the waitlist for specialists is long, and patients may miss a critical window for early interventions or mitigations of risk factors for AD. Therefore, the solution lies in bridging this diagnostic gap at the primary care level by developing an easily administered and interpretable method to screen for AD risk and involving behavioral health consultants (BHCs), typically clinical psychologists, to carry out the screening and collaborate with PCPs to mitigate the risk factors.

The advancement of machine learning models offers a vast avenue for aiding the diagnostic process due to their speed and data-driven decisions that often excel in comparison to humans.^3^ Recent efforts to develop machine learning models to assist clinicians in identifying early-stage AD have demonstrated robust accuracy.^4,5^ However, the use of these models has raised important issues pertaining to the needs for inaccessible data and interpretability due to their “black box” nature in generating the output, potentially leading to a lack of trust in the outputs from clinicians.^6,7^ Interpretable machine learning models (IML; i.e., white-box approach), on the other hand, do not suffer from the same issues as it provides the “why” of outputs, offering insights into how specific features contribute to predictions and allowing for transparent and understandable decision-making processes, which has been shown to promote trust between clinicians and the machine learning outputs.^8^

In this study, we developed risk scores that were presented in a scorecard model to assess the risk of developing AD using the FasterRisk algorithm. Risk scores are predictive models that have been used in various fields, including medicine, to aid decision-making processes through basic mathematical calculation.^9–11^ The FasterRisk algorithm is a recent advancement that significantly improves the creation of high-quality risk scores that exceeds the performance of traditional methods, such as rounding logistic regression coefficients or non-data-driven approaches.^12^ We selected the following variables to develop the scorecards due to their practicability and comprehensive representation of factors influencing AD: demographic information, cognitive tests from various domains, and daily functioning. We designed the scorecards to help inform clinicians of the probable risk of developing AD based on a patient’s presentation, aiding in decisions about when to refer patients to specialists and involving BHCs to initiate interventions while the patients are on the waitlist for specialists. We predicted that our framework could generate a scoring system with robust predictive power using accessible variables.

## Materials and methods

### Participants

We included data from 676 baseline visits from several Alzheimer’s Disease Neuroimaging Initiative database (ADNI 1, 2, GO, and 3; adni.loni.usc.edu) as of August 2023 and 4592 baseline visits from the National Alzheimer’s Coordinating Center (NACC) Uniform Data Set (UDS) extracted in March 2024. ADNI was launched in 2003 as a public-private partnership, led by Principal Investigator Michael W. Weiner, MD. The primary goal of ADNI has been to test whether serial magnetic resonance imaging (MRI), positron emission tomography (PET), other biological markers, and clinical and neuropsychological assessment can be combined to measure the progression of mild cognitive impairment (MCI) and early AD.

ADNI’s broader criteria include age 55-90, a minimum of 6 years of education, consistent medication for the past 4 weeks, Hachinski scale < 4 (to rule out vascular dementia), and Geriatric Depression Scale < 6; more information can be found www.adni-info.org. We conducted performance validation of the model using an independent cohort, NACC UDS, which is a multi-site study that has been collecting data from Alzheimer’s Disease Research Centers (ADRCs) since 2005. In general, NACC UDS inclusion criteria for enrollment include an absence of neurological disorders, severe mental illness, learning disability, and substance abuse. The data consists of participants’ demographics, cognitive status, cognitive performance, and other clinical history and information; more information can be found www.naccdata.org.

We included participants in our analysis who were classified by ADNI and NACC as having normal cognition (NC) or amnestic MCI. Participants were divided into two groups: stable and progressive. The stable group consisted of individuals who remained at the same diagnosis level over time. The progressive group included those who developed AD at later visits. Specifically, participants who progressed from aMCI to AD or NC to AD were placed in the progressive group. To be included in the stable group, the sample had to contain data indicating at least 3 years of aMCI and 8 years of NC. Individuals who reverted to aMCI from NC were not included in the analysis. We excluded participants with non-amnestic MCI or those who developed other types of dementia. Participants with invalid or missing values were identified and removed from both ADNI (n = 11; 1.63%) and NACC (n = 1219; 25%).

### Neuropsychological Tests and Functioning

We selected a range of neuropsychological tests that tapped into a variety of cognitive domains, such as attention, executive function, memory (short-term and long-term), and language/verbal fluency. The selected tests were the Logical Memory Immediate (LIMM), Logical Memory Delayed (LDEL), Category Animal (CATANIMSC), Trail Making Test A (TMT A), and Trail Making Test B (TMT B). These tests were selected because they were administered across all ADNI and NACC cohorts. Additionally, we included the instrumental activities of daily living measured with the Functional Activities Questionnaire (FAQ). Some adjustments were made for the NACC dataset because some tests were replaced or not administered. For example, the later NACC cohorts (UDS v.3) completed Craft Story 21 instead of Logical Memory (UDS v.2). We converted the test scores to match Logical Memory based on the conversion table provided by NACC crosswalk study.^13^

### Data Preprocessing

To prepare the data for analysis, we converted categorical variables into numerical representations through Scikit-learn Labelencoder. The next preprocessing step was applying binarization using the FasterRisk built-in binarization module to convert the features from continuous into binary features (Figure 1). This ensures the proper input data format for the algorithm.

**Figure 1.**
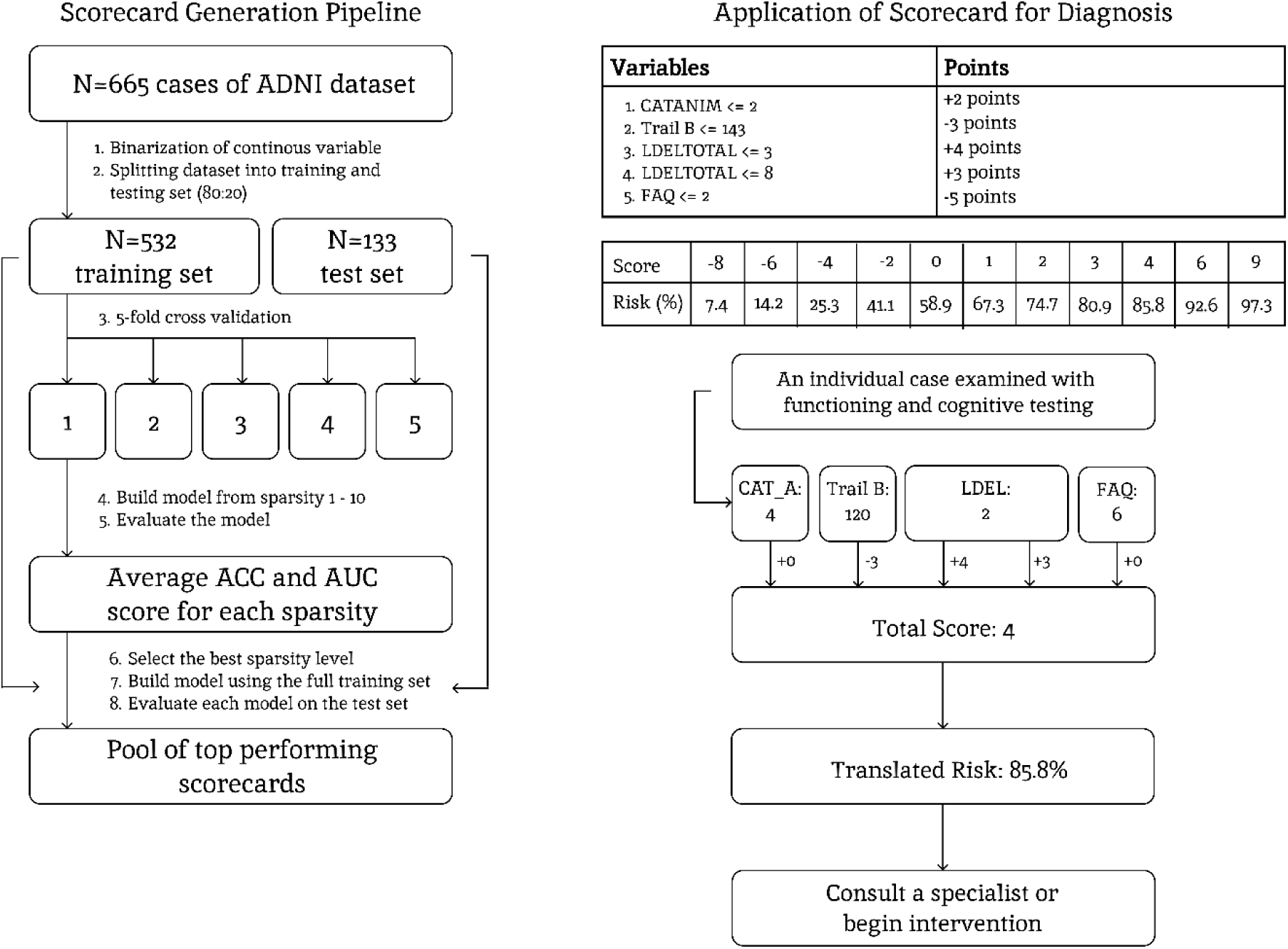
Pipeline of conducting FasterRisk algorithm to generate the FLAME scorecard and its clinical application.

### Risk Scorecard Generation and Optimization

We employed the FasterRisk algorithm to develop an interpretable, point-based risk scorecard, a model type widely used for transparent clinical decision-making.^12^ Specific risk factors are assigned simple, whole-number points (e.g., +5 points for a given condition). A clinician sums up these points to calculate a patient’s total risk score, which corresponds to a risk percentage, making each decision transparent and easy to explain. The algorithm then constructs risk scores through an iterative process. It starts by identifying the single most predictive risk factor to form a base model. Then, it incrementally adds new factors one by one, keeping only those that improve the model’s performance. The algorithm also explores other combinations by swapping existing features with other promising candidates. This approach was selected because it significantly outperforms RiskSlim, a previous state-of-the-art model for finding risk scores.^14^ The scorecard’s complexity is controlled by a ‘sparsity’ parameter, k, which dictates the exact number of predictive features included. To find the optimal balance between simplicity and accuracy, we systematically tested models with 1 to 10 features using 5-fold cross-validation (Figure 1).

### Final Model Evaluation

The final scorecard was trained on 80% of ADNI data and its performance was internally validated on the held-out 20% test set (Figure 1). To ensure the stability of our results, we generated and analyzed the top 10 scorecards produced by the algorithm. By doing this, we were able to examine the set of most consistently predictive features. The model’s performance was evaluated using accuracy, where a 50% risk threshold determined classification, and also the Area Under the Receiver Operating Curve (AUC), which quantifies the model’s overall ability to discriminate between classes. We validated the best scorecard out of the 10 generated scorecards on NACC dataset. All computations were performed on Python 3.12.3 and data preprocessing was done using Numpy 1.23.5.

## Results

### Participant Characteristics

We included data from 665 ADNI participants (164 NC and 501 aMCI) and 3657 NACC participants (2846 NC and 810 aMCI) from 41 ADRCs at baseline visits. Over time, 43.8% (*n* = 291) of ADNI and 33.1% (*n* = 1212) of NACC participants were diagnosed with AD. As shown in Table 1, ADNI participants were 44.1% female, with a mean age of 73.44 years and an average of 16.15 years of education, and NACC participants were 65.80% female, with a mean age of 73.44 years and an average of 16.15 years of education. There were statistically significant group differences between stable and progressive in ADNI (age at *p* < 0.01 and education at *p* < 0.05) and NACC (age and education at *p* < 0.001). The average transitions for the aMCI-AD and the NC-AD groups are 1.09 years and 7.10 years for ADNI and 3.02 years and 7.20 years for NACC.

**Table 1.** Table of ADNI and NACC demographic, cognition, and functioning by groups.

| Variables (ADNI) | Stable (*n* = 374) | Progressive (*n* = 291) | All (*n* = 665) |
| --- | --- | --- | --- |
| Age (years) | 72.8 (6.74) ** | 74.27 (7.03) ** | 73.44 (6.90) |
| Education (years) | 16.34 (2.72) * | 15.89 (2.82) * | 16.15 (2.77) |
| Female (%) | 169 (45.2%) | 124 (42.6%) | 293 (44.1%) |
| Race (White%) | 350 (93.6%) | 274 (94.2%) | 624 (93.83%) |
| FAQ | 0.98 (2.56) *** | 4.90 (5.00) *** | 2.70 (4.29) |
| Category Animal | 19.91 (5.43) *** | 16.11 (4.94) *** | 18.25 (5.55) |
| TMT A | 35.29 (14.42) *** | 45.9 (24.2) *** | 39.93 (20.01) |
| TMT B | 87.91 (46.57) *** | 134.63 (73.83) *** | 108.36 (64.32) |
| LM immediate | 11.66 (3.84) *** | 7.63 (3.88) *** | 9.90 (4.35) |
| LM delayed | 9.83 (4.21) *** | 4.59 (4.13) *** | 7.54 (4.92) |
| Variables (NACC) | Stable (*n* = 2445) | Progressive (*n* = 1212) | All (*n* = 3657) |
| Age (years) | 68.16 (9.55) *** | 76.61 (8.49) *** | 70.96 (10.03) |
| Education (years) | 16.61 (6.15) *** | 15.85 (5.6) *** | 16.36 (5.98) |
| Female (%) | 1646 (67.3%) | 759 (62.6%) | 2405 (65.8%) |
| Race (White%) | 2074 (84.8%) | 1075 (88.7%) | 3149 (86.11%) |
| FAQ | 0.26 (1.33) *** | 2.59 (4.66) *** | 1.03 (3.1) |
| Category Animal | 21.87 (5.66) *** | 17.70 (5.66) *** | 20.49 (5.99) |
| TMT B | 80.35 (79.88) *** | 140.73 (168.42) *** | 100.36 (120.29) |
| LM delayed | 12.91 (4.19) *** | 7.68 (5.26) *** | 11.18 (5.19) |

ADNI = Alzheimer's Disease Neuroimaging Initiative; NACC = National Alzheimer's Coordinating Center; FAQ = Functional Activities Questionnaire; TMT A = Trail Making Test A; TMT B = Trail Making Test B; LM = Logical Memory
Differences between stable and progressive, \* $p < 0.05$ , \*\* $p < 0.01$ , \*\*\* $p < 0.001$ □ □
FAQ was adjusted for age.

### Group Differences in Functioning and Cognitive Performance at Baseline

We compared group differences in cognitive performance and daily functioning using analysis of covariance (ANCOVA) between stable and progressive groups. The FAQ was adjusted for age, and cognitive measures were adjusted for age and education. All measures showed statistical differences between stable and progressive groups in both ADNI and NACC (*p* < 0.001). The progressive groups showed lower performance and scores in verbal fluency/language, TMT A, TMT B, immediate and delayed memory, and daily functioning. Higher values on TMT A and TMT B indicate worse performance.

### Alzheimer Prediction Risk Score

Based on the FasterRisk algorithm, a sparsity level of 5 was selected for the most optimal combination for the generation of the final scorecards to predict AD development. Ten scorecards were generated with a test AUC ranging from 0.868 to 0.892 and accuracy between 81.20% and 84.21% (Supplementary Figure 1). The scorecard with the highest test AUC (0.892) shown in Table 2 represents Category Animal ≤ 20 (2 points), Trail Making Test B ≤ 143 (−3 points), Logical Memory Delayed ≤ 3 (4 points), Logical Memory Delayed ≤ 8 (3 points), and FAQ ≤ 2 (−5 points). Positive points contribute to increased total score of the scorecard, while negative points decrease it. The total score corresponds to the risk percentage shown in Table 2. The probable AD development risk was 7.4% for a total score of −8, 19.1% for a score of −5, 32.7% for a score of −3, 50% for a score of −1, 74.7% for a score of 2, 85.8% for a score of 4, and greater than 90% for a score of 6, 7, or 9 (Table 2). We refer to this scorecard model as the FLAME scorecard, a mnemonic representing functioning, language, memory, and executive function. We validated this scorecard with NACC dataset, yielding a test AUC of 0.795, suggesting good generalizability of the model.

**Table 2.** Scorecard with the highest AUC and Risk Score to assess AD development probability.

| Variables | Points |  |
| --- | --- | --- |
| 1. Category Animal $\leq 20$ | + 2 points | ... |
| 2. Trail Making Test B $\leq 143$ | -3 points | + ... |
| 3. Logical Memory Delayed $\leq 3$ | + 4 points | + ... |
| 4. Logical Memory Delayed $\leq 8$ | + 3 points | + ... |
| 5. FAQ $\leq 2$ | -5 points | + |
| **SCORE** |  | = |

LDEL = Logical Memory delayed recall; Test; □FAQ=Functional Activities Questionnaire

| Score | -8 | -6 | -4 | -3 | -1 | 1 | 2 | 4 | 6 | 9 |
| --- | --- | --- | --- | --- | --- | --- | --- | --- | --- | --- |
| Risk (%) | 7.4 | 14.2 | 25.3 | 32.7 | 50.0 | 67.3 | 74.7 | 85.8 | 92.6 | 97.3 |

## Discussion

Our study presents a novel approach to predicting the risk of developing AD that offers potential to be applied in integrated primary care settings by employing a set of obtainable variables, including demographics, daily functioning, and cognitive performance. By utilizing the FasterRisk algorithm, we generated ten scorecards, each achieved high predictive accuracy (81.20%–84.21%) and strong discriminative performance (AUC = 0.868–0.892), reflecting a robust balance between sensitivity and specificity in identifying individuals at risk of developing AD. We further validated one of the ten scorecards on a separate cohort, indicating good generalizability. Additionally, the FLAME scorecard compared favorably to established machine learning models for predicting AD development,^15,16^ and compared similarly or slightly below previously studied interpretable machine learning models incorporating biological (AUC = 0.86) and neuroimaging data (accuracy = 87.5%).^17,18^

A key advantage of the FLAME scorecard lies in its potential clinical practicality at integrated primary care settings. We selected variables that can be implemented and fit in one appointment with a behavior health consultant (BHC), which typically lasts approximately 30 minutes. The BHC would also have sufficient time to gather brief information related to patients’ concerns and general mental wellbeing. Following this assessment, the BHCs can input the results into the scorecard and collaborate with the PCPs to determine appropriate next steps, which may vary depending on the findings and presenting concerns. For instance, the PCPs may refer patients to neurologists or other specialists, start a pharmacological treatment to slow down memory decline (e.g., acetylcholinesterase inhibitors), reduce the use of anticholinergic medications, order p-tau 217 blood test, or recommend patients to make follow-up appointments with the BHCs to address behavioral factors that could mitigate memory decline (e.g., sleep hygiene, engaging in physical activity, medication adherence, mood symptoms). BHCs may also address safety concerns (e.g., driving, living arrangement) and make appropriate recommendations. These services could serve as valuable interim interventions while patients are placed on the waitlist for specialists.

Another advantage of the FLAME scorecard lies in its interpretability, which provides clinicians with a clear explanation of the specific features influencing its predictions. This transparency promotes human-computer interaction, in this case, trust between clinicians and the machine learning outputs.^8^ Furthermore, these scorecards offer flexibility in their implementation, which allows clinicians to incorporate their expertise into the scorecards when predicting the risk of AD development. For example, on the FLAME scorecard, having a FAQ score of 2 or below would decrease the total by 5 points. If a patient indicates that the reason for requiring assistance on some activities is due to physical limitations rather than decreased mental capacity, then the clinician can consider this context when evaluating the predicted risk of AD. The balance between performance, interpretability, and flexibility positions our scorecard as a promising tool for practical clinical application, where understanding the rationale behind predictions is paramount for effective and informed decision support.

There are some limitations in our study. The scorecards generated in this study are only applicable to typical AD with an amnestic profile. Recent reviews highlighted that a subset of AD populations presents an atypical profile, such as deficits in executive functioning, language, visuospatial, and other non-memory domains, and this subset is typically younger than those who develop typical AD^19,20^. This scorecard might also be limited to discern typical AD presentation from limbic predominant age-related TDP-43 encephalopathy (LATE) due to similarity in their cognitive profiles.^21,22^ Future studies including participants with atypical profiles or LATE are necessary to better inform PCPs of which follow-up tests to order to help confirm their diagnosis, which will cut down some costs compared to sending the patients to all tests/procedures.

Additionally, the demographic composition of the ADNI and NACC samples, predominantly White and highly educated individuals, highlights the need for further validation in more diverse populations to ensure the generalizability of our findings. Lastly, it was beyond the scope of our study to determine a definitive cutoff value for clinical decision-making.

Our study lays the groundwork for a more accessible and population-wide approach to screening for Alzheimer’s disease. Moving forward, we aim to collaborate with primary care physicians and behavioral health consultants to collect both qualitative and quantitative data on the feasibility and potential impact of implementing these scorecards in routine clinical practice. This collaboration will provide valuable insights into the practical challenges and opportunities for integrating our tool into the healthcare system.

## Conclusions

Our study generated a robust scoring system for predicting the likelihood of developing Alzheimer’s disease using accessible and cost-efficient variables through interpretable machine learning. This framework’s interpretability may aid clinicians in integrated primary care settings in providing early detection to their patients, including those residing in resource-constrained areas.

## Data Availability

All data produced are available online at https://adni.loni.usc.edu and https://naccdata.org

## Acknowledgments

Data collection and sharing for this project was funded by the Alzheimer’s Disease Neuroimaging Initiative (ADNI) (National Institutes of Health Grant U01 AG024904) and DOD ADNI (Department of Defense award number W81XWH-12-2-0012). ADNI is funded by the National Institute on Aging, the National Institute of Biomedical Imaging and Bioengineering, and through generous contributions from the following: AbbVie, Alzheimer’s Association; Alzheimer’s Drug Discovery Foundation; Araclon Biotech; BioClinica, Inc.; Biogen; Bristol-Myers Squibb Company; CereSpir, Inc.; Cogstate; Eisai Inc.; Elan Pharmaceuticals, Inc.; Eli Lilly and Company; EuroImmun; F. Hoffmann-La Roche Ltd and its affiliated company Genentech, Inc.; Fujirebio; GE Healthcare; IXICO Ltd.; Janssen Alzheimer Immunotherapy Research & Development, LLC.; Johnson & Johnson Pharmaceutical Research & Development LLC.; Lumosity; Lundbeck; Merck & Co., Inc.; Meso Scale Diagnostics, LLC.; NeuroRx Research; Neurotrack Technologies; Novartis Pharmaceuticals Corporation; Pfizer Inc.; Piramal Imaging; Servier; Takeda Pharmaceutical Company; and Transition Therapeutics. The Canadian Institutes of Health Research is providing funds to support ADNI clinical sites in Canada. Private sector contributions are facilitated by the Foundation for the National Institutes of Health (www.fnih.org). The grantee organization is the Northern California Institute for Research and Education, and the study is coordinated by the Alzheimer’s Therapeutic Research Institute at the University of Southern California. ADNI data are disseminated by the Laboratory for Neuro Imaging at the University of Southern California.

The NACC database is funded by NIA/NIH Grant U24 AG072122. NACC data are contributed by the NIA-funded ADRCs: P30 AG062429 (PI James Brewer, MD, PhD), P30 AG066468 (PI Oscar Lopez, MD), P30 AG062421 (PI Bradley Hyman, MD, PhD), P30 AG066509 (PI Thomas Grabowski, MD), P30 AG066514 (PI Mary Sano, PhD), P30 AG066530 (PI Helena Chui, MD), P30 AG066507 (PI Marilyn Albert, PhD), P30 AG066444 (PI David Holtzman, MD), P30 AG066518 (PI Lisa Silbert, MD, MCR), P30 AG066512 (PI Thomas Wisniewski, MD), P30 AG066462 (PI Scott Small, MD), P30 AG072979 (PI David Wolk, MD), P30 AG072972 (PI Charles DeCarli, MD), P30 AG072976 (PI Andrew Saykin, PsyD), P30 AG072975 (PI Julie A. Schneider, MD, MS), P30 AG072978 (PI Ann McKee, MD), P30 AG072977 (PI Robert Vassar, PhD), P30 AG066519 (PI Frank LaFerla, PhD), P30 AG062677 (PI Ronald Petersen, MD, PhD), P30 AG079280 (PI Jessica Langbaum, PhD), P30 AG062422 (PI Gil Rabinovici, MD), P30 AG066511 (PI Allan Levey, MD, PhD), P30 AG072946 (PI Linda Van Eldik, PhD), P30 AG062715 (PI Sanjay Asthana, MD, FRCP), P30 AG072973 (PI Russell Swerdlow, MD), P30 AG066506 (PI Glenn Smith, PhD, ABPP), P30 AG066508 (PI Stephen Strittmatter, MD, PhD), P30 AG066515 (PI Victor Henderson, MD, MS), P30 AG072947 (PI Suzanne Craft, PhD), P30 AG072931 (PI Henry Paulson, MD, PhD), P30 AG066546 (PI Sudha Seshadri, MD), P30 AG086401 (PI Erik Roberson, MD, PhD), P30 AG086404 (PI Gary Rosenberg, MD), P20 AG068082 (PI Angela Jefferson, PhD), P30 AG072958 (PI Heather Whitson, MD), P30 AG072959 (PI James Leverenz, MD). AW was supported by an NIH grant from NIGMS and OD 1P20GM152280.

