## Supplemental file for "Development of Alzheimer’s Disease Risk Score for Future Primary Care: A White-Box Approach"

**Scorecard 1**

| Variables | Points |
| --- | --- |
| 1. Category Animal ≤ 21 2. Trail Making Test B ≤ 143 3. Logical Memory Delayed ≤ 3 4. Logical Memory Delayed ≤ 8 5. FAQ ≤ 2 | +2 points  -3 points  +4 points  +2 points  -4 points |

Optimized risk score for AD development probability of Scorecard 1

| Score | -7 | -5 | -3 | -2 | -1 | 0 | 2 | 3 | 5 | 8 |
| --- | --- | --- | --- | --- | --- | --- | --- | --- | --- | --- |
| Risk (%) | 7.0 | 15.2 | 29.7 | 39.4 | 50.0 | 60.6 | 78.4 | 84.8 | 93.0 | 98.0 |

The training accuracy and AUC are 79.699% and 0.871

The test accuracy and AUC are 81.203% and 0.885

**Scorecard 2**

| Variables | Points |
| --- | --- |
| 1. Category Animal ≤ 21 2. Trail Making Test B ≤ 140 3. Logical Memory Delayed ≤ 3 4. Logical Memory Delayed ≤ 8 5. FAQ ≤ 2 | +2 points  -3 points  +4 points  +2 points  -4 points |

Optimized risk score for AD development probability of Scorecard 2

| Score | -7 | -5 | -3 | -2 | -1 | 0 | 2 | 3 | 5 | 8 |
| --- | --- | --- | --- | --- | --- | --- | --- | --- | --- | --- |
| Risk (%) | 7.0 | 15.1 | 29.6 | 39.4 | 50.0 | 60.6 | 78.5 | 84.9 | 93.0 | 98.0 |

The training accuracy and AUC are 79.699% and 0.870

The test accuracy and AUC are 81.203% and 0.885

**Scorecard 3**

| Variables | Points |
| --- | --- |
| 1. Category Animal ≤ 21 2. Trail Making Test B ≤ 144 3. Logical Memory Delayed ≤ 3 4. Logical Memory Delayed ≤ 8 5. FAQ ≤ 2 | +2 points  -3 points  +4 points  +2 points  -4 points |

Optimized risk score for AD development probability of Scorecard 3

| Score | -7 | -5 | -3 | -2 | -1 | 0 | 2 | 3 | 5 | 8 |
| --- | --- | --- | --- | --- | --- | --- | --- | --- | --- | --- |
| Risk (%) | 7.0 | 15.1 | 29.7 | 39.4 | 50.0 | 60.6 | 78.5 | 84.9 | 93.0 | 98.0 |

The training accuracy and AUC are 79.699% and 0.870

The test accuracy and AUC are 81.203% and 0.885

**Scorecard 4**

| Variables | Points |
| --- | --- |
| 1. Category Animal ≤ 21 2. Trail Making Test B ≤ 141 3. Logical Memory Delayed ≤ 3 4. Logical Memory Delayed ≤ 8 5. FAQ ≤ 2 | +2 points  -3 points  +4 points  +2 points  -4 points |

Optimized risk score for AD development probability of Scorecard 4

| Score | -7 | -5 | -3 | -2 | -1 | 0 | 2 | 3 | 5 | 8 |
| --- | --- | --- | --- | --- | --- | --- | --- | --- | --- | --- |
| Risk (%) | 7.0 | 15.1 | 29.7 | 39.4 | 50.0 | 60.6 | 78.5 | 84.9 | 93.0 | 98.0 |

The training accuracy and AUC are 79.699% and 0.870

The test accuracy and AUC are 81.203% and 0.885

**Scorecard 5**

| Variables | Points |
| --- | --- |
| 1. Category Animal ≤ 21 2. Trail Making Test B ≤ 137 3. Logical Memory Delayed ≤ 3 4. Logical Memory Delayed ≤ 8 5. FAQ ≤ 2 | +3 points  -3 points  +4 points  +3 points  -5 points |

Optimized risk score for AD development probability of Scorecard 5

| Score | -8 | -5 | -3 | -2 | 0 | 2 | 3 | 5 | 7 | 10 |
| --- | --- | --- | --- | --- | --- | --- | --- | --- | --- | --- |
| Risk (%) | 5.3 | 14.2 | 25.3 | 32.7 | 50.0 | 67.3 | 74.7 | 85.8 | 92.6 | 97.3 |

The training accuracy and AUC are 79.699% and 0.869

The test accuracy and AUC are 81.203% and 0.886

**Scorecard 6**

| Variables | Points |
| --- | --- |
| 1. Category Animal ≤ 21 2. Trail Making Test B ≤ 136 3. Logical Memory Delayed ≤ 3 4. Logical Memory Delayed ≤ 8 5. FAQ ≤ 2 | +3 points  -3 points  +4 points  +3 points  -5 points |

Optimized risk score for AD development probability of Scorecard 6

| Score | -8 | -5 | -3 | -2 | 0 | 2 | 3 | 5 | 7 | 10 |
| --- | --- | --- | --- | --- | --- | --- | --- | --- | --- | --- |
| Risk (%) | 5.3 | 14.2 | 25.3 | 32.7 | 50.0 | 67.3 | 74.7 | 85.8 | 92.6 | 97.3 |

The training accuracy and AUC are 79.699% and 0.869

The test accuracy and AUC are 81.203% and 0.886

**Scorecard 7**

| Variables | Points |
| --- | --- |
| 1. Category Animal ≤ 21 2. Trail Making Test B ≤ 148 3. Logical Memory Delayed ≤ 3 4. Logical Memory Delayed ≤ 8 5. FAQ ≤ 2 | +2 points  -3 points  +4 points  +2 points  -4 points |

Optimized risk score for AD development probability of Scorecard 7

| Score | -7 | -5 | -3 | -2 | -1 | 0 | 2 | 3 | 5 | 8 |
| --- | --- | --- | --- | --- | --- | --- | --- | --- | --- | --- |
| Risk (%) | 7.0 | 15.2 | 29.7 | 39.4 | 50.0 | 60.6 | 78.4 | 84.8 | 93.0 | 98.0 |

The training accuracy and AUC are 79.699% and 0.870

The test accuracy and AUC are 81.203% and 0.885

**Scorecard 8**

| Variables | Points |
| --- | --- |
| 1. Category Animal ≤ 21 2. Trail Making Test B ≤ 143 3. Logical Memory Delayed ≤ 1 4. Logical Memory Delayed ≤ 8 5. FAQ ≤ 2 | +2 points  -3 points  +4 points  +3 points  -4 points |

Optimized risk score for AD development probability of Scorecard 8

| Score | -7 | -5 | -3 | -2 | -1 | 0 | 2 | 3 | 5 | 8 |
| --- | --- | --- | --- | --- | --- | --- | --- | --- | --- | --- |
| Risk (%) | 6.8 | 14.9 | 29.5 | 39.3 | 50.0 | 60.7 | 78.7 | 85.1 | 93.2 | 98.7 |

The training accuracy and AUC are 80.639% and 0.870

The test accuracy and AUC are 83.459% and 0.868

**Scorecard 9**

| Variables | Points |
| --- | --- |
| 1. Category Animal ≤ 21 2. Trail Making Test B ≤ 180 3. Logical Memory Delayed ≤ 3 4. Logical Memory Delayed ≤ 8 5. FAQ ≤ 2 | +3 points  -4 points  +4 points  +3 points  -5 points |

Optimized risk score for AD development probability of Scorecard 9

| Score | -9 | -6 | -4 | -3 | -1 | 1 | 2 | 4 | 6 | 10 |
| --- | --- | --- | --- | --- | --- | --- | --- | --- | --- | --- |
| Risk (%) | 5.3 | 14.1 | 25.3 | 32.7 | 50.0 | 67.3 | 74.7 | 85.9 | 92.6 | 98.2 |

The training accuracy and AUC are 80.075% and 0.869

The test accuracy and AUC are 81.203% and 0.884
